# Atorvastatin and standard treatment of *Helicobacter pylori*: Randomized Clinical Trial

**DOI:** 10.1101/2020.05.29.20117200

**Authors:** Mohammad Reza Mohammad Hoseini Azar, Parham Portaghali, Amin Sedokani, Ali Jafari

## Abstract

**Background:** Considering the increase in drug resistance over time to *Helicobacter pylori* treatment relying on the anti-inflammatory and antibacterial effects of atorvastatin to increase the success rate of *H. pylori* eradication, we examined the effect of adding atorvastatin to standard treatment of *H. pylori* eradication.

**Materials and Methods:** A total of 186 symptomatic patients who had been diagnosed with *Helicobacter pylori* infection and tested for *H. pylori* eradication were examined by a pathological response or positive urea breath test. Patients who received atorvastatin in addition to standard treatment were also identified based on a table of random numbers. Standard treatment included a 240^mg^ bismuth subcitrate tablet, a 40^mg^ pantoprazole tablet, a 500^mg^ metronidazole tablet, and 2 capsules of 500^mg^ amoxicillin, all taken BID for 14 days. After 4 weeks of treatment, all patients underwent stool testing for *H. pylori* fecal antigen. If the test was positive, the request was considered a failure of treatment, and if the test was negative, it was considered a successful eradication of *H. pylori*. The clinical trial registration code for this study is IRCT20190823044589N1.

**Results:** The eradication rate of *H. pylori* was 80% in the control group and 80.9% in the intervention group, which did not show a statistically significant difference between the two groups (P-value=0.971).

**Conclusion:** Adding atorvastatin to 4-drug regimen of PPI, bismuth subcitrate, amoxicillin, and metronidazole as the first line of treatment for *H. pylori* eradication is ineffective.

**Significance of this Study (Summary box):**

- **What is already known about this subject?**
  1. Infection of *H. pylori* is common worldwide and the antibiotic resistance is increasing
  2. Atorvastatin, has anti-inflammatory and antibacterial effects. But also, have D grade interaction with clarithromycin in *H. pylori* eradication regimen and increases the toxic and lethal risk of atorvastatin toxicity.
- **What are the new findings?**
  1. Eradication rate of *H. pylori* using the standard treatment of a 240^mg^ bismuth subcitrate tablet, a 40^mg^ pantoprazole tablet, a 500^mg^ metronidazole tablet, and 2 capsules of 500^mg^ amoxicillin, BID for 14 days, is at least 80%.
  2. Adding atorvastatin to 4-drug regimen of PPI, bismuth subcitrate, amoxicillin, and metronidazole as the first line of treatment for *H. pylori* eradication is ineffective.
- **How might it impact on clinical practice in the foreseeable future?**
  1. There should be more analysis on cost-benefice of adding atorvastatin to standard regimen of treatment for *H. pylori* eradication, but adding the atorvastatin to metronidazole-based *H. pylori* treatment is ineffective and for clarithromycin-based treatment is dangerous.

## Introduction

*Helicobacter pylori* is a spiral gram-negative bacterium that clones in the human stomach and is one of the most important pathogens known (1-3). Infection with this bacterium is common around the world, and some studies have shown that about half of the world’s population is infected with the bacterium (4). In developed countries, the infection with this bacterium is decreasing, but in developing countries, the rate of infection with this bacterium is still high (2, 4, 5). Studies in the Middle East have reported very different numbers, however, the prevalence of *H. pylori* infection in the Middle East appears to be close to 60% or more (2). Numerous studies have been conducted in Iran, most of which reported a prevalence of more than 60%, which is almost similar to other developing countries in the Middle East (2, 6). *H. pylori* cause inflammation in the stomach and duodenum, including gastric ulcer disease, duodenal ulcer disease, iron deficiency, and vitamin B_12_ deficiency, gastric adenocarcinoma, and gastric ulcer, and MALT (B cell mucosa-associated lymphoid tissue lymphoma) (7, 8). Studies have shown an association between *H. pylori* infection and diseases such as autoimmune diseases, cardiovascular disease, and metabolic syndrome (6, 9-11). Studies have also shown that extra-gastric diseases associated with H. pylori are associated with higher levels of serum LDL (12). Therefore, eradication of *H. pylori* has been accepted as a treatment strategy in gastric lymphoma and gastric ulcer and as a strategy for the prevention of gastric adenocarcinoma (7, 8). Also, its eradication is being studied in the treatment of metabolic syndrome and cardiovascular disease (10, 13). The first line of treatment for *H. pylori* eradication based on various guidelines around the world is the use of a proton pump inhibitor (PPI) and two antimicrobial agents (clarithromycin and amoxicillin/metronidazole). The success rate of *H. pylori* eradication in previous studies was reported to be approximately 80% to 96%, but in recent studies this rate has been reported to be below 80%. Much of this reduction in effect is due to drug resistance to antibiotics, and the rate of drug resistance increases over time(3, 14). Therefore, finding alternative therapies is needed to improve the eradication of Helicobacter pylori.

Statins, including simvastatin and atorvastatin, has been used as one of the main therapies for hyperlipidemia and is used in patients with ischemic heart disease (IHD) for primary and secondary prophylaxis (15, 16). One of the mechanisms of statin drugs in improving the prognosis of patients with IHD and atherosclerosis is the reduction of CRP (C-Reactive Protein), which is known as an acute phase reactant (17). Also, studies have shown antibacterial and anti-inflammatory effects in this class of drugs (18-20). Also, studies have shown that pravastatin has an anti-inflammatory effect on gastritis, and taking any statin-class medication at any time significantly reduces the risk of gastric cancer (21, 22).

Due to the anti-inflammatory and antibacterial effects of statins and the fact that studies have shown that *H. pylori* increase cytokine production and inflammatory factors (23), statins have been suggested as one of the drugs that help eliminate Helicobacter pylori. In-vivo studies have shown that pravastatin reduces the incidence of leukocyte infiltration and the severity of gastritis caused by Helicobacter pylori, but does not affect stability. However, pravastatin has only been fed to mice for 7 days. And the effects of longer-term use have not been investigated (22). On the other hand, in a clinical study, it was stated that the administration of simvastatin for one week as an adjunct drug helps to eliminate *H. pylori* by treating osteoarthritis (3). Also, in laboratory studies, the high-dose anti-inflammatory effects of atorvastatin have been reported to be greater than those of simvastatin (24). Overall, given the aforementioned discussions and studies on the effect of statins on the elimination of Helicobacter pylori, relying on the anti-inflammatory and antibacterial effects of atorvastatin to increase the success rate of *H. pylori* eradication, the effect of adding atorvastatin to Standard treatment for *H. pylori* eradication.

## Material and Methods

The present study is a single-blinded randomized parallel clinical trial gastrointestinal ward of Imam Hospital of Urmia University of Medical Sciences has been performed. The study included 186 symptomatic patients who had been diagnosed with *H. pylori* infection by a pathological response or positive respiratory urea test and were candidates for *H. pylori* eradication candidate. Take a history of all patients before entering the study. The criteria for admission are as follows: The patient enters the study. For patients who had the conditions to enter the study, after explaining the plan, such as the importance of the study and the benefits of the study, the study method and the fact that the patient is able to leave the study at any time and the cost of all actions is the responsibility of the research team. If the patient was satisfied with the participation in the study, after obtaining written consent from the patient, they entered the study.

According to the table of random numbers, patients who received atorvastatin in addition to standard treatment were identified. After the patient entered the study, standard treatment was prescribed for the patient, and patients who were randomly assigned to receive atorvastatin based on a table of random numbers were also given a pack of 14 panel drugs. Necessary explanations on how to take the medication were explained to the patient by the treating physician. Patient characteristics including name, gender and age of the patient were recorded. Standard treatment includes a 240^mg^ bismuth subcitrate tablet, a 40^mg^ pantoprazole tablet, a 500^mg^ metronidazole tablet, and 2 capsules of 500^mg^ amoxicillin, all taken BID^1^ for 14 days. The adjuvant treatment with atorvastatin was 40^mg^ and was placed in a drug pack every 24 hours for 14 days. After the end of the treatment period, if more than 85% of the medications were taken, it was acceptable and the patient was considered to be adhering to the treatment, otherwise the non-adherence to the treatment was considered and the patient was excluded from the study. After 4 weeks of treatment, all patients underwent a stool test to check for *H. pylori* fecal antigen. If the test was positive, the result was recorded as a failure of treatment, and if the test was negative, it was recorded as a successful elimination of *H. pylori*. The patient was also asked about the possible side effects of the medication, which would be recorded if any side effects occurred. In case of any possible side effects of atorvastatin (such as muscle disorders, muscle cramps, abnormal fatigue, diarrhea, arthralgia, myalgia), the patient’s medication was discontinued and recorded.

### Inclusion criteria

All of enrolled patients must had the following criteria: Age ≥18 y/o, lack of liver disease at the same time, lack of diabetes at the same time, not taking statins in the last 6 months, no history of gastric surgery, no history of *H. pylori* eradication treatment, not take antibiotics, PPI, histamine (H_2_) receptor blockers, anti-inflammatory drugs, bismuth salts in the past month, Lack of allergy to any of the antibiotics used in the study, lack of gastrointestinal malignancy, lack of active gastrointestinal bleeding, non-pregnant and non-breastfeeding, and no history of radiation therapy.

For patients who had the conditions to enter the study, after explaining the plan, such as the importance of the study and the benefits of the study, the study method and the fact that the patient is able to leave the study at any time and the cost of all actions is the responsibility of the research team. If the patient was satisfied with the participation in the study, after obtaining written consent from the patient, they entered the study.

### Data Analysis

All the information obtained from the present study was entered in SPSS V. 26 software. Qualitative data were reported as frequency and percentage and quantitative data as mean±standard deviation. To compare the frequency of elimination of *H. pylori* between the two groups and also based on sex in each group, chi-square test and based on age in each group, independent t-test was used. Significant levels in the tests were considered ≤0.05. All statistical tests were analyzed by a statistician, and after the statistical process, the results were announced by a member of the project who was not involved in selecting the type of treatment for patients and statistical analysis and was aware of the distribution of drugs in drug packages.

### Ethical Statement

After the approval of the university’s ethics committee (<u>IR.UMSU.REC</u>), the plan was implemented without imposing additional costs on patients and informed consent obtained from each patient. The study group will be published and the individual results will be presented if necessary, without mentioning the name and personal details. Ethical approval is under code of <u>IR.UMSU.REC.1398.186</u> and the clinical trial registration code is <u>IRCT20190823044589N1</u>.

## Results

In the present study, 93 patients in each treatment group and a total of 186 patients were enrolled. 3 patients of control group and 4 patients from the intervention group were excluded due to lack of follow-up the study. In the control group, out of 90 patients, 48 were male and 42 were female, and out of 89 patients in the intervention group, 50 were male and 39 were female patients, there was no statistically significant difference between the two groups (p=0.38). The mean age of patients in the control group was 54.6±6.1 years and in the intervention group was 57.4±3.8 years, which did not show a statistically significant difference between the two groups (p=0.73).

### Eradication Rate

Of the 90 control patients who underwent *H. pylori* eradication, in in 72 (80%) patients, the test was negative (successful eradication) and in18 (20%) had a positive test that indicated treatment failure. Of these 18 patients, 11 (61%) were male and 7 (39%) were female. Of the 89 patients in the intervention group who were examined for *H. pylori* eradication, 72 (80.8%) patients were reported to be negative and 17 (19.2%) patients reported treatment failure, of which 9 were male and 8 were female. Statistically, between the two groups, the treatment failure rate of the intervention group was 19.1% and the control group was 20%, which did not show a statistically significant difference between the two groups (p-value=0.97). Also, there was no significant statistically significant difference between patients with treatment failure (p-value=0.62).

### Age

The mean age of patients with treatment failure in the control group and intervention was 59.6±8.3 and 59.4±5.7 years, respectively, but no significant statistical difference was observed between the two groups (p-value=0.98).

## Discussion

The first line of treatment for *H. pylori* eradication based on various guidelines around the world is the use of a proton pump inhibitor (PPI) and two antimicrobial agents (clarithromycin, amoxicillin or metronidazole). The success rate of *H. pylori* eradication in previous studies was reported to be about 80% to 96%, but in recent studies this rate has been reported below 80% (3). Much of this reduction in effect is due to drug resistance to antibiotics and the rate of drug resistance increases over time (3, 14). Therefore, finding alternative therapies is needed to improve the eradication of *H. pylori*.

As a general rule, the physician should choose a medication regimen for the patient that has a 90% eradication rate (25). According to studies conducted in Iran, one of the first treatment regimens for *H. pylori* is 4-drug regimen of PPI, bismuth, amoxicillin and metronidazole or clarithromycin (26). However, studies show that over time, the success rate of these drug regimens in eradicating *H. pylori* has decreased from 90% to about 80% (26) and even in some studies it has decreased to less than 70% (27). Therefore, adding adjuvant medications or changing diet therapy seems to be necessary to eradicate *H. pylori*.

Therefore, due to the anti-inflammatory and antibacterial effects of statins and the fact that studies have shown that *H. pylori* increase cytokine production and inflammatory factors (23), statins are one of the drugs that can help eliminate *H. pylori*. In the present study, due to the severe drug interactions of clarithromycin and atorvastatin (D class interaction) (28, 29) for the standard treatment regimen of a 240^mg^ bismuth subcitrate tablet, a 40^mg^ pantoprazole tablet, a 500^mg^ metronidazole tablet and 2 capsules of 500^mg^ amoxicillin, all of which were used BID for 14 days, and in the intervention group, in addition to receiving a standard diet, patients were also prescribed 40^mg^ of atorvastatin. In the present study, the eradication rate of *H. pylori* was 80% in the control group and 80.9% in the intervention group, which did not show a statistically significant difference between the two groups (p-value=0.97), while in the study of Nseir et al, and another study by Shakerian et al, it was found that adding statins to a 4-drug regimen significantly increased the success rate of *H. pylori* eradication. In a 2012 study by Nseir et al, in the placebo group, the rate of elimination of *H. pylori* was 72%, and for those receiving simvastatin, in addition to the standard treatment, it was 91%, with a statistically significant difference of p-value of 0.03 (3).In the Shakerian study, 110 patients in the control group received a 14-day diet of amoxicillin, clarithromycin, bismuth, and esomeprazole, and 110 patients in the day-to-day intervention group received 40^mg^ of atorvastatin with an antibiotic regimen for 14 days. The test results were tested a month later using *H. pylori* fecal antigen testing. *H. pylori* eradication rates in the intervention and control groups were 78.18% and 65.45%, respectively (p-value=0.025) (30). Comparing the two studies with the present study, it is found that adding statins to the 4-drug regimen containing clarithromycin increases the eradication rate of *H. pylori*, while in the therapeutic regimen used in the present study, clarithromycin was replaced by metronidazole, and adding statins does not statistically improve *H. pylori* eradication.

Studies have shown that clarithromycin inhibits CATP3A4 by inhibiting OATP1B1 and OATP1B3 cells in hepatocyte cells, which can lead to complications such as AV block, rhabdomyolysis, acute renal failure, hyperkalemia, and death (31-37). Therefore, the positive effect of statins on regimen containing clarithromycin may be due to the high accumulation of statins, which on the one hand increases the eradication rate of *H. pylori* eradication, but on the other hand will have dangerous side effects. Therefore, the combination of this class of drugs with clarithromycin in the anti-Helicobacter pylori diet does not appear to be beneficial. On the other hand, it should be noted that the drug regimen used in the study of Shakerian et al, the is not recommended regimen in Iran (study place). However, the success rate of a treatment regimen is related to geographical area, age and sex, and as reported in the results of their study, the intervention and control groups were 78.18% and 65.65%, respectively (30). Which was less than the results of the present study (80.9% and 80%). Therefore, in addition to avoiding the use of statin drugs at the same time as clarithromycin, it is recommended that statins be studied with other preferred regimens in the region.

## Data Availability

The data of this study is available to the professional reviewers.

## Footnotes

1 bis in die, Twice a day

